# A single-nucleus transcriptomic atlas of human basal ganglia during development forwarding diagnosis and therapy of pediatric movement disorders

**DOI:** 10.64898/2026.06.04.26354648

**Authors:** Birthe Katrin Alexandra Lange, Eugenio Graceffo, Werner Stenzel, Heike Biebermann, Markus Schuelke, Nina-Maria Wilpert

## Abstract

Gene therapy is rapidly emerging as a transformative treatment for monogenic neurological disorders, including pediatric movement disorders such as aromatic L-amino acid decarboxylase (AADC) deficiency. However, its success critically depends on defining target cells and windows for therapeutic intervention. Here, we present an open-access single-nucleus transcriptomic atlas of the human basal ganglia spanning a therapy-relevant window from second/third trimester to the perinatal period and adulthood. Across 35,755 nuclei, we identify major (non-)neuronal cell types, retrace developmental trajectories, and characterize gene-regulatory networks. We identify so far unrecognized human-specific expression of key neuronal signaling genes, including *GNAO1* and *ADCY5*, and discuss the implications for targeted gene replacement therapies. Unexpectedly, we found that the Huntingtin gene (*HTT*) is already expressed during prenatal stages of human brain development, supporting a previously proposed neurodevelopmental component of Huntington’s disease, which should be considered in diagnostic and therapeutic strategies. Moreover, *FOXG1* expression and regulon activity are predominantly located in a prenatal time window, suggesting constraints on the effectiveness of postnatal interventions. Our findings highlight the importance of datasets capturing human brain development in real time and provide a publicly available resource to guide precision gene therapy strategies in the future.

**Graphical abstract:** 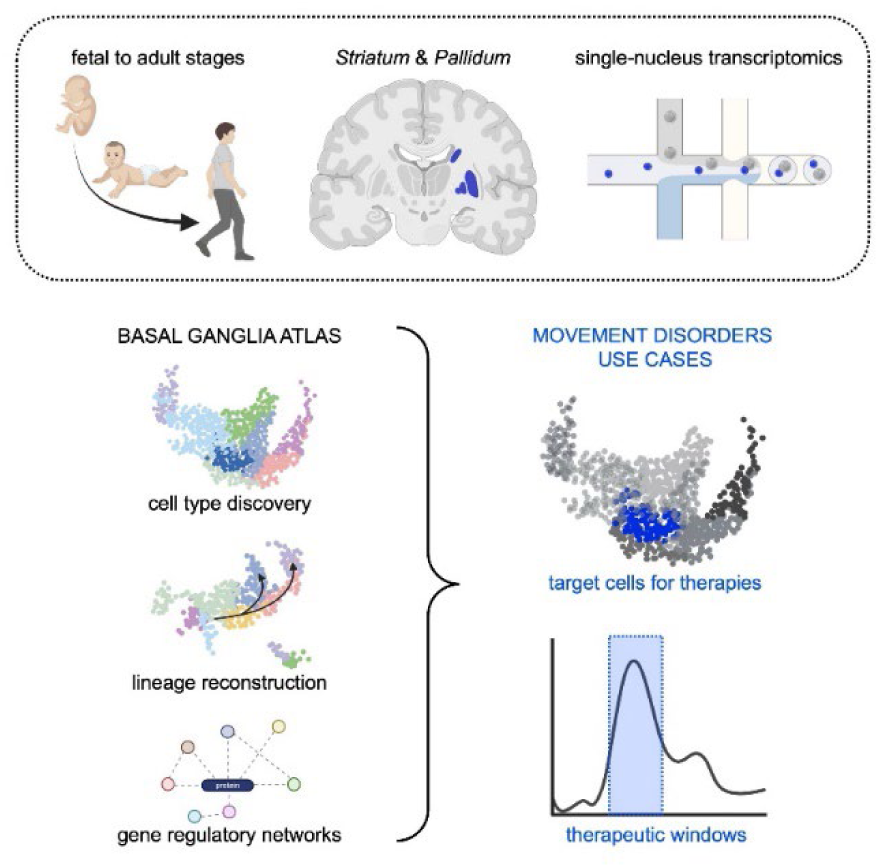

## Introduction

After decades of research, gene replacement therapy has now become a viable treatment option for genetic disorders. A growing number of gene therapies have received regulatory approval, and the development pipeline continues to expand exponentially. While the U.S. Food and Drug Administration (FDA) projected in 2019 that 10-20 gene therapy products could be approved annually by 2025,^1^ a recent report from the American Society of Gene and Cell Therapy shows that 41 gene therapies were approved worldwide in the final quarter of 2025 alone,^2^ underscoring the rapid acceleration of the field. These approaches hold transformative potential, offering the possibility to alter the course of a disease and reduce the long-term healthcare burden.^3^

Pediatric neurology is a field that has long been limited largely to supportive treatments. However, increasing rates of genetic diagnoses driven by the growing availability of routine genetic testing in many countries,^4^ are now enabling the development of promising gene therapies. A striking example is the adeno-associated virus 2 (AAV2)-based gene therapy for aromatic L-amino acid decarboxylase (AADC) deficiency (OMIM# 608643), an autosomal recessive disorder caused by dysfunction of AADC, the enzyme catalyzing the final step in dopamine and serotonin synthesis. Untreated patients typically present with severe neurodevelopmental delay and a complex movement disorder characterized by hypokinesia, dystonia, painful oculogyric crises, and autonomic dysfunction.^5^ For this and other therapies that have to be specifically delivered into the brain parenchyma, both the target region (delivery to the *Putamen*^6^ or the ventral tegmental area^7^ in the case of AADC deficiency) and the timing of administration are critically important. Clinical studies have demonstrated remarkable therapeutic effects of the gene therapy for AADC deficiency, provided the treatment is administered early, with children may improve from absent head control to independent walking.^8^ However, this example also highlights the risk of potential “over-therapy”, as many treated patients develop dyskinesias, particularly when the therapy is initiated late.^8,9^ One proposed mechanism for this side effect could be an increased sensitivity to the newly restored dopaminergic signaling circuits, *e.g.* through a prior compensatory upregulation of dopamine receptor density. Taken together, these observations underscore the importance of understanding the physiological spatiotemporal expression of disease-associated genes and their pathway components in order to define appropriate target cell populations and optimal therapeutic windows.

Here, we focus on the disease group of pediatric movement disorders, whose pathophysiology is localized to the basal ganglia, a set of subcortical nuclei. The basal ganglia comprise several interconnected structures, including the *Striatum* (consisting of the *Putamen* and the *Nucleus caudatus*), the *Globus pallidus*, and the *Nucleus accumbens*. In contrast to the cerebral cortex, these regions comprise predominantly inhibitory neurons, including projection neurons and interneurons, that release gamma-aminobutyric acid (GABA) as their main neurotransmitter. These inhibitory neurons originate from the transient embryonic ganglionic eminences (GE) and migrate to their target regions in the basal ganglia and cortical areas during early to late gestation. The basal ganglia are central components of cortico-basal ganglia-thalamo-cortical circuits that support motor control, limbic processing, and learning. Their impairment causes movement disorders and psychiatric disease.^10^

Recent years have seen major advances in our understanding of the development and function of human cortical areas.^11–13^ In contrast, knowledge of developing subcortical regions remains comparatively limited. Until recently, only a small number of studies had explored the diversity of GE and GE-derived cells across development and species, including embryonic^14,15^ and adult human,^16^ non-human primates,^17^ and mice,^18^ primarily using single-cell transcriptomic approaches. Very recently, in early 2026, four preprints were released in rapid succession, highlighting the growing interest in the molecular characterization of the human basal ganglia.^19–22^ Heffel *et al.* presented a comprehensive atlas integrating single-nucleus DNA methylation and 3D chromatin architecture (snm3C-seq) with multiplexed single-cell spatial transcriptomics (limited to 6,175 transcripts) across basal ganglia tissue from mid-gestation to adulthood.^19^ Shortly thereafter, Zhang *et al.* introduced a platform for the collection of single-cell and epigenomic datasets of the basal ganglia.^20^ This was followed by two additional pre- prints providing single-cell transcriptomic data on adult basal ganglia tissue.^21,22^ Despite these advances, a comprehensive single-nucleus RNA sequencing (snRNA-seq) analysis of the developing human basal ganglia covering second trimester to perinatal stages was still lacking, particularly with respect to the expression of movement disorder-associated genes. The second and third trimester and the early infantile period are characterized by rapid acquisition of motor function and representing a critical window for therapeutic interventions.

Here, we address this knowledge gap by generating a single-nucleus transcriptomic atlas of human basal ganglia across pre- and postnatal development. Our open-access resource will assist the rational definition of therapeutic windows and of target cell populations, thereby supporting the development of targeted gene therapies for pediatric movement disorders.

## Results

### Major cell classes in the developing human basal ganglia

We generated a single-nucleus transcriptional atlas of human basal ganglia development by performing snRNA-seq on samples from seven donors obtained from the NIH NeuroBioBank (fetal and perinatal cases) and one from the Department of Neuropathology at the Charité – University Medical Center Berlin (adult case; **Figure 1A**). Tissue samples included the *Striatum*, in most cases the *Globus pallidus*, and in some cases the *Nucleus accumbens* (**Figure 1B**, **Table 1**). We focused our analysis on the second to third trimester and the perinatal period. This time window is crucial for the development of motor capabilities with children rapidly acquiring motor milestones; from the inability to perform voluntary movements to achieving head control, grasping, and rolling over. This period may represent a critical therapeutic window to apply early gene therapy for pediatric movement disorders.^6,7^ Such early interventions could become possible with the help of genetic newborn screening,^23^ which is presently being tested in pilot projects.^24^

**Figure 1.**
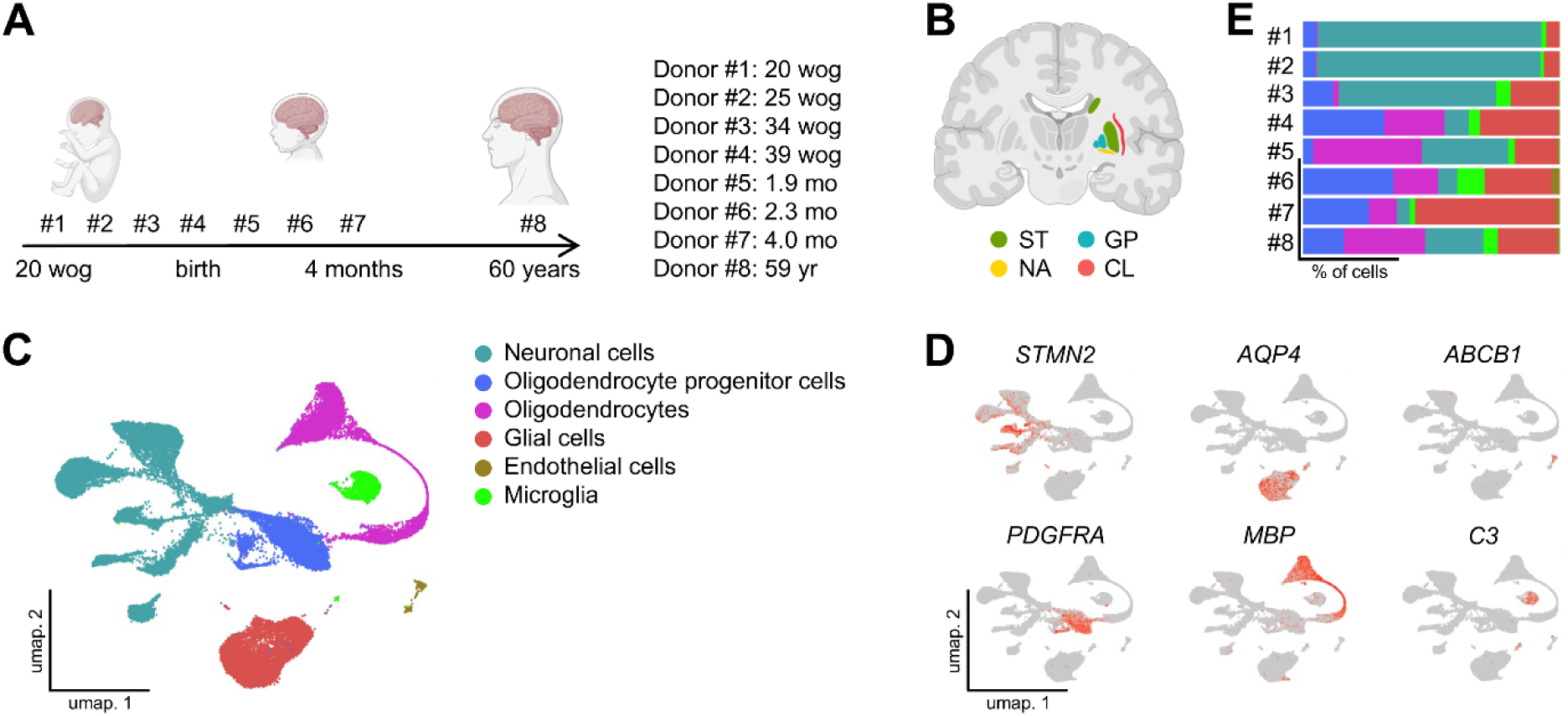
Cell type taxonomy of the developing human basal ganglia. **(A)** Schematic of the developing brain and subject ages included in this study; wog, week of gestation; mo, month; yr, year. **(B)** Color-coded representation of the included basal ganglia and adjacent regions: ST, *Striatum*; GP, *Globus pallidus*; NA, *Nucleus accumbens*; CL, *Claustrum*. **(C)** UMAP (uniform manifold approximation and projection) plot depicting 35,755 nuclei assigned to six major clusters. **(D)** Expression of marker genes used for the annotation of major cell-types. **(E)** Fraction of each cell type in total cells from each individual donor. Color code according to (C).

**Table 1.** Clinical and neuropathological characteristics of brain tissue donors. Donor samples #1–7 were obtained from the NIH NeuroBioBank, and sample #8 from the Department of Neuropathology at Charité – University Medical Center Berlin. Samples #1–7 were classified as control tissue by the NIH NeuroBioBank, as there were no neurological co-morbidities and the causes of death were not primarily neurological; this also applied to sample #8. The *Putamen* was available from all donors, whereas the *Nucleus caudatus*, *Globus pallidus*, and *Nucleus accumbens* were available only in selected cases, as indicated. M, male; F, female; na, not assessed.

| Donor # | Age<br>[month] | Gestational<br>age [week] | Postnatal<br>survival<br>[day] | Post mortem<br>delay [hour] | Sex | *Putamen* | *Nucleus*<br>*caudatus* | *Globus*<br>*pallidus* | *Nucleus ac-*<br>*cumbens* | Cause of death |
| --- | --- | --- | --- | --- | --- | --- | --- | --- | --- | --- |
| 1 | 0 | 20 | 0 | 16 | M | x |  | x |  | prematurity, retroplacental hematoma |
| 2 | 0 | 25 | 1 | 16 | F | x | x | x |  | complications of prematurity |
| 3 | 0 | 34 | 0 | 23 | F | x | x |  |  | hypoplastic left heart syndrome |
| 4 | 0 | 39 | 1 | 17 | F | x | x | x |  | multi organ failure |
| 5 | 1.9 | 40 | 56 | 11 | M | x |  | x |  | congenital heart defect |
| 6 | 2.3 | 40 | 69 | 27 | M | x | x | x | x | asphyxia |
| 7 | 4.0 | 40 | 120 | 23 | M | x | x |  | x | pneumonia |
| 8 | ~60<br>years | na | na | 20 | F | x | x |  |  | pneumonia, respiratory insufficiency |

Brain tissue was obtained from donors, whose developmental stages spanned prenatal (20– 39 gestational weeks), early postnatal (1.9–4.0 months), and adult (∼60 years) stages (**Figure 1A**). Nuclei were clustered according to their gene expression profiles in a hypothesis-free manner, resolving the major neuronal and non-neuronal cell types (**Figure 1C**). Feature plots depicting the expression of canonical markers used for cell type annotation are shown in **Figure 1D**. Each sample contained cells from all major neuronal and non-neuronal cell classes, supporting consistent anatomical targeting across the intended regions and developmental stages (**Figure 1E, Figure S1A–H**). In prenatal samples, neuronal populations were predominant relative to other cell types, while non-neuronal populations, such as oligodendrocyte (progenitor) cells, increased markedly around birth, which is consistent with the perinatal proliferation of oligodendrocytes and the progression of myelination. Altogether, we generated an integrated dataset of 35,755 nuclei capturing the development of the *Striatum* and *Pallidum*, ranging from mid-gestational progenitors to mature adult neurons.

### Transcriptional diversity of developing neuronal subtypes

In the process, we focused on a more detailed characterization of those neuronal subpopulations that represent cell types primarily involved in movement disorders. To resolve the neuronal heterogeneity, we performed sub-clustering of the nuclei and detected transcriptional diversity reflecting their brain regions of origin (**Figures 2A-B**). The majority of nuclei originated from the *Striatum*, consistent with the fact that the *Striatum* was the only brain region available across all donors (**Table 1**). *Globus pallidus*-derived nuclei segregated into two discrete clusters, in line with their known embryonic origins from the *PENK*-positive lateral GE (LGE) and *NKX2.1*-positive medial GE (MGE), with their embryonic origin emerging as the primary determinant of transcriptional proximity, overriding later regional identity (**Figure 2A, 2C**). Unexpectedly, in addition to the predominant GABAergic cells of the basal ganglia, we detected a glutamatergic neuronal cluster (*SATB2*^+^), which transcriptionally corresponded to the anatomically adjacent tissue of the *Claustrum* (*GNG2^+^, NTNG2^+^, GNB4^+^*; **Figure 1B, 2A-B**, data not shown).^25^

**Figure 2.**
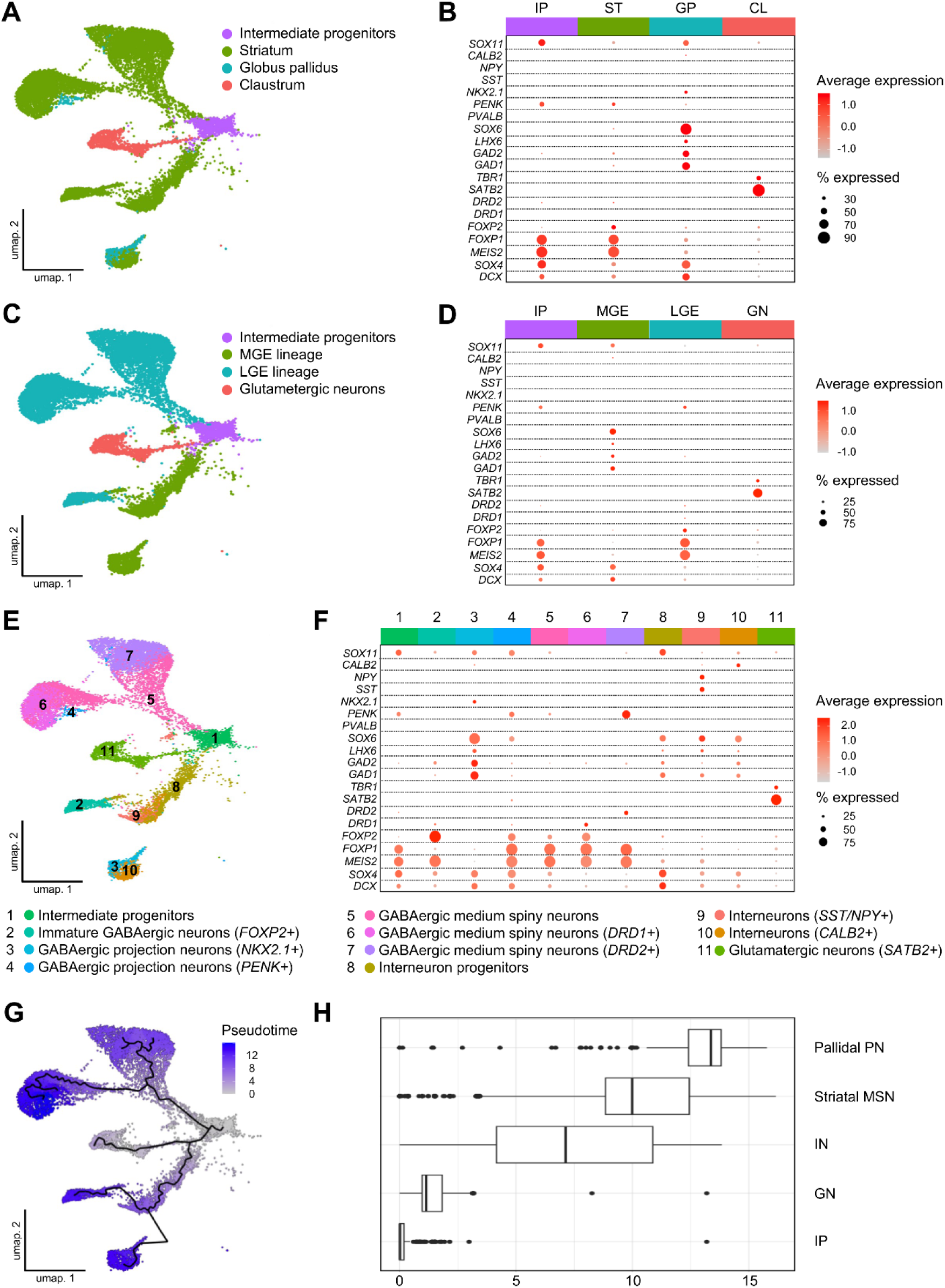
Characterization of neuronal subtypes. (A, C, **E)** UMAP plots of neuronal nuclei from the snRNA-seq dataset classified according to **(A)** originating brain regions, **(C)** embryonic origin, and **(E)** fine neuronal subtypes. **(B, D, F)** Dot plots of marker genes used for cell-type annotations. **(B)** IP, intermediate progenitors; ST, *Striatum*; GP, *Globus pallidus*; CL, *Claustrum*. **(D)** IP, intermediate progenitors; MGE, medial ganglionic eminence; LGE, lateral ganglionic eminence; GN, glutamatergic neurons. **(G)** trajectories of neurons according to the Monocle3 software. **(H)** Distributions of single-nucleus Pseudotime pseudotime scores for neuronal subtypes. High scores reflect larger transcriptional differences from the cluster of origin. PN, projection neurons; MSN, medium spiny neurons; IN, interneurons; GN, glutamatergic neurons; IP, intermediate progenitors.

GEs are transient structures of the prenatal brain that give rise to GABAergic neurons and inhibitory interneurons, which subsequently migrate toward their subcortical and cortical destinations.^26^ With the help of our dataset, we confirmed that striatal GABAergic medium spiny neurons (MSN) and *PENK*- and *ISL1*-double-positive pallidal GABAergic projection neurons originate from the LGE (**Figures 2C-F**). In contrast, striatal interneurons and *NKX2.1*-positive pallidal GABAergic projection neurons were derived from the MGE (**Figures 2C-F**).^18^

Our fine-grained cell-type annotation identified the largest neuronal population as striatal MSN, comprising dopamine receptor D1 (*DRD1*)-positive (direct pathway, which facilitates movement), *DRD2*-positive (indirect-pathway, which suppresses movement), and double-negative subpopulations (**Figure 2E-F**).^14,17,19,21,22^ Although recent studies have proposed a further subdivision into matrix-MSN, which give rise to the canonical direct and indirect pathways, and striosome-MSN, which project to midbrain dopaminergic neurons, as well as *DRD1*/*DRD2* co-expressing ‘eccentric’ MSN,^18,19^ these populations did not robustly demarcate in our dataset, likely due to the limited number of neuronal nuclei and snRNA-seq data sparsity, which makes it difficult to detect rare cell populations. We identified an LGE-derived GABAergic neuronal cluster (**Figure 2E-F**, cluster 2) that could not be confidently assigned to either striatal MSN or pallidal GABAergic projection neurons. Based on the expression of *DCX*, *SOX4*, and *MEIS2*, marking intermediate progenitor-like states, together with high *FOXP2* expression, a transcription factor implicated in MSN differentiation,^27^ we annotated this population as ‘immature GABAergic neurons’. Additional interneuron subtypes expressing *SST/NPY* or *CALB2* emerged as discrete populations.^19^

We performed pseudotime analysis to characterize neuronal maturation trajectories and identified three major trajectories originating from intermediate progenitors (**Figure 2G-H**). Based on these trajectories, the *DRD1*- and *DRD2*-double-negative MSN subpopulation may represent MSN progenitors.

Together, our findings define a spectrum of neuronal subtypes that integrates regional identity, developmental origin, and maturation state within the striatal-pallidal system.

### Spatiotemporal expression of genes relevant for the diagnosis of movement disorders

As exome and genome sequencing are progressively integrated into routine diagnostic processes, we are confronted with a growing number of variants that have to be interpreted regarding their disease-causing potential. In the field of movement disorders this had led to a surge of newly described disease entities.^28^ At the same time, gene therapies are rapidly being developed for local^7^ or systemic^29,30^ application. For a maximal effect of such therapies, it is essential to know the cell types and developmental time windows, that a defective gene is active in. Since the Movement Disorder Society Genetic Mutation Database (MDSGene)^31^ continuously curates a list of genes that are involved in movement disorders, we concentrated on these genes for our further analysis; especially on those associated with an early manifestation age (examples are shown in **Figure 3**).

**Figure 3.**
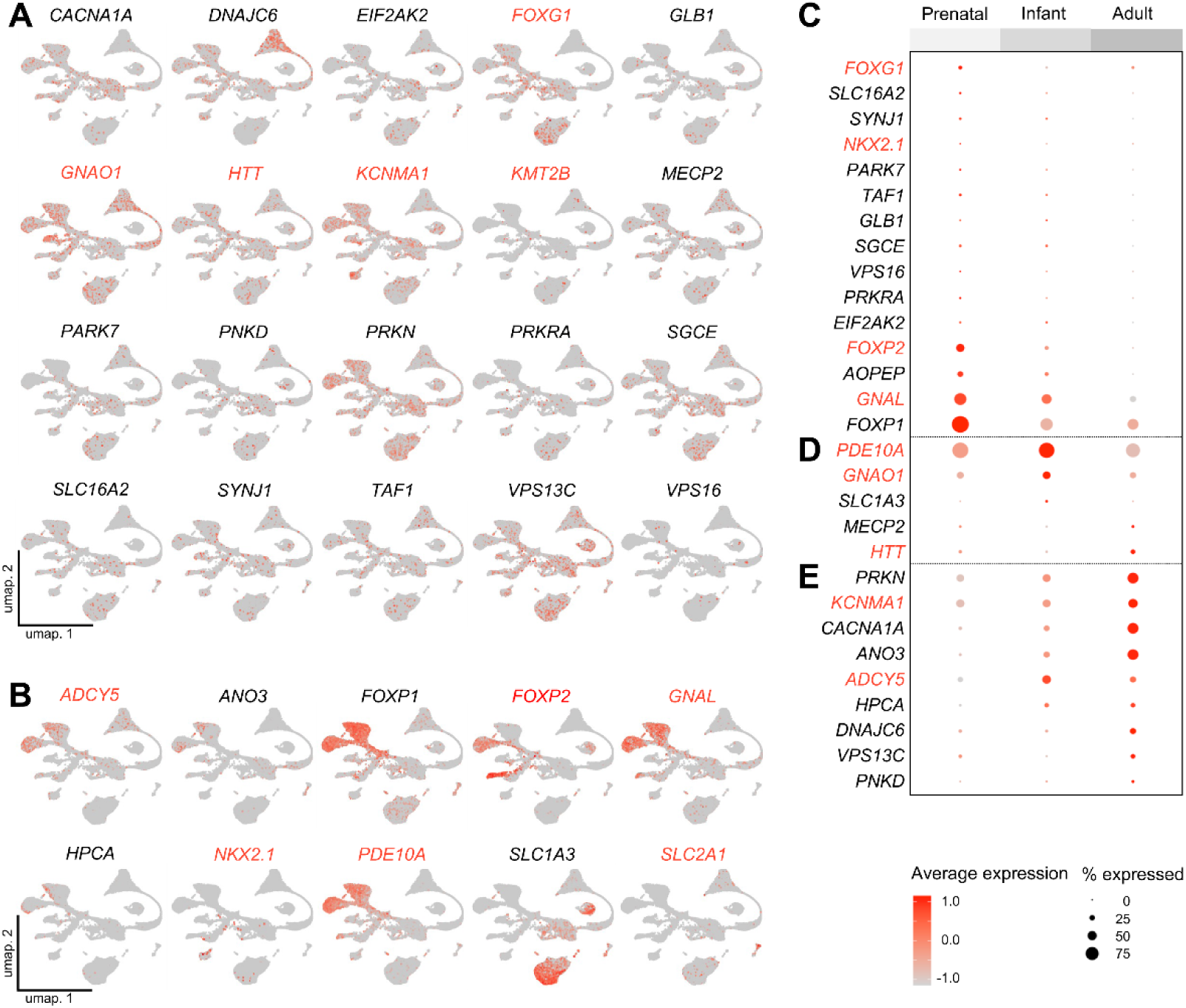
Spatiotemporal expression of movement disorder-related genes. (A, **B)** UMAP plots colored by expression of the respective movement-disorder relevant genes, listed in the MDSGene database.^31^ Genes were categorized based on their expression patterns into expressed **(A)** in the majority of cell types analyzed *versus* **(B)** in a cell type-specific fashion. **(C–E)** Dot plots depicting the stratification of selected genes according to their temporal dynamics into genes predominantly expressed prenatally **(C)**, expressed throughout development **(D)**, or showing increasing expression postnatally **(E)**. Genes discussed in the text are highlighted in red.

We classified the genes into groups of ‘general’ (**Figure 3A**) and ‘cell type-specific’ expression patterns (**Figure 3B**) and report them in alphabetical order. Postsynaptic dopaminergic signaling action in the basal ganglia involves the G protein-coupled receptors (GPCR) and cyclic adenosine monophosphate (cAMP) cascade.^32^ *ADCY5, GNAL, GNAO1*, and *PDE10A* encode key components thereof, such as adenylate cyclases, subunits of GPCR, and phosphodiesterases. In our dataset, *GNAO1* expression was not restricted to the *Striatum*, the current primary target region of intraparenchymal gene therapy approaches,^33^ but showed ubiquitous expression in non-neuronal and neuronal cells and was also detected in nuclei originating from the *Globus pallidus*. The solute carrier *SLC2A1*, on the other hand, showed predominant expression in endothelial cells,^34^ while the adenylate cyclase *ADCY5* expression was enriched in MSN. Notably, in contrast to previous reports on rodents,^35^ *ADCY5* expression in our dataset was not limited to *DRD1*-positive MSN but was similarly detected in *DRD2*-positive MSN.

We next stratified pediatric movement disorder-associated genes according to the temporal window of their predominant expression, ranging from ‘primarily prenatal’ (**Figure 3C**) *via* ‘throughout development’ (**Figure 3D**) to ‘predominantly postnatal expression’ (**Figure 3E**). We grouped samples #1–4 (**Table 1**), ranging from the second trimester to birth, under the term ‘prenatal’. The ‘infantile’ stage included samples #5–7, spanning the postnatal period up to 4 months of age, while the ‘adult’ stage was represented by sample #8. For stratification purposes, we compared gene expression exclusively in MSN of the *Striatum*, as this was the only neuronal cell type available across all developmental stages. As an example, we want to highlight the transcription factor FOXG1, which is a key regulator of forebrain development.^36^ *FOXG1* is mutated in the FOXG1 syndrome, which is characterized by a severe neurodevelopmental and movement disorder, for which postnatal gene therapy approaches are currently being developed.^29^ In our dataset, *FOXG1* exhibited predominantly prenatal and only minimal postnatal expression (**Figure 3C**). In contrast, we observed that *huntingtin* (*HTT*), classically associated with adult-onset neurodegeneration in Huntington’s disease (HD), showed already substantial expression in prenatal brain samples (**Figure 3D**). Genes primarily implicated in neuronal signaling and function, rather than in early neurodevelopment, such as the adenylate cyclase *ADCY5*, the phosphodiesterase *PDE10A*, and the potassium channel *KCNMA1* displayed increasing expression toward ‘infantile’ stages and were detectable in more transcriptionally distinct cells, *i.e.* potentially more differentiated cells, along the pseudotime trajectories (**Figure 2G**), underscoring a high potential for successful postnatal therapeutic interventions in patients carrying pathogenic variants in these genes (**Figure 3D-E**).^37,38^

### Cell type-specific gene-regulatory networks in the basal ganglia

Transcript levels alone do not necessarily reflect protein abundance and activity, therefore providing only limited insight into the time windows, during which therapeutic interventions may be most effective. Therefore, we applied the program package SCENIC to infer the activity of transcription factors or other regulators of gene expression by looking on the expression of their downstream targets (‘regulons’).^39^ The analysis of the transcription factor NKX2.1 shall serve as an example. NKX2.1 is crucial for basal ganglia development and mutations cause disease phenotypes that comprise childhood onset chorea (*e.g.* benign hereditary chorea, OMIM #118700 or choreoathetosis, hypothyroidism and neonatal respiratory distress, OMIM #610978). Its detected expression levels were minimal (**Figure 3B-C**), whereas its regulon activity, reflecting its activity as a transcription factor on downstream targets, was high to very high in pallidal GABAergic projection neurons and striatal interneurons (**Figure 4A**)^40^ confirming its relevant biological activity. A similar pattern was observed for the methyltransferase *KMT2B*, which was expressed at very low levels (**Figure 3A**) but showed high regulon activity in intermediate progenitors (**Figure 4A**). The gene encodes a lysine methyltransferase that activates transcription *via* methylation of histone 3 lysine 4 (H3K4). Mutations cause childhood onset dystonia type 28 (OMIM #617284). In contrast, *FOXG1* displayed not only markedly reduced expression levels after birth (**Figure 3C**), but even a further decline of regulon activity in successive postnatal samples (#5–8 in **Figure 4B-C**), indicating continuously diminishing transcriptional activity after birth, potentially curtailing the effect of postnatal therapeutic interventions. FOXP2 and THRB, on the other hand, two known regulators of neuronal maturation, remained active postnatally (**Figure 4B**).

**Figure 4.**
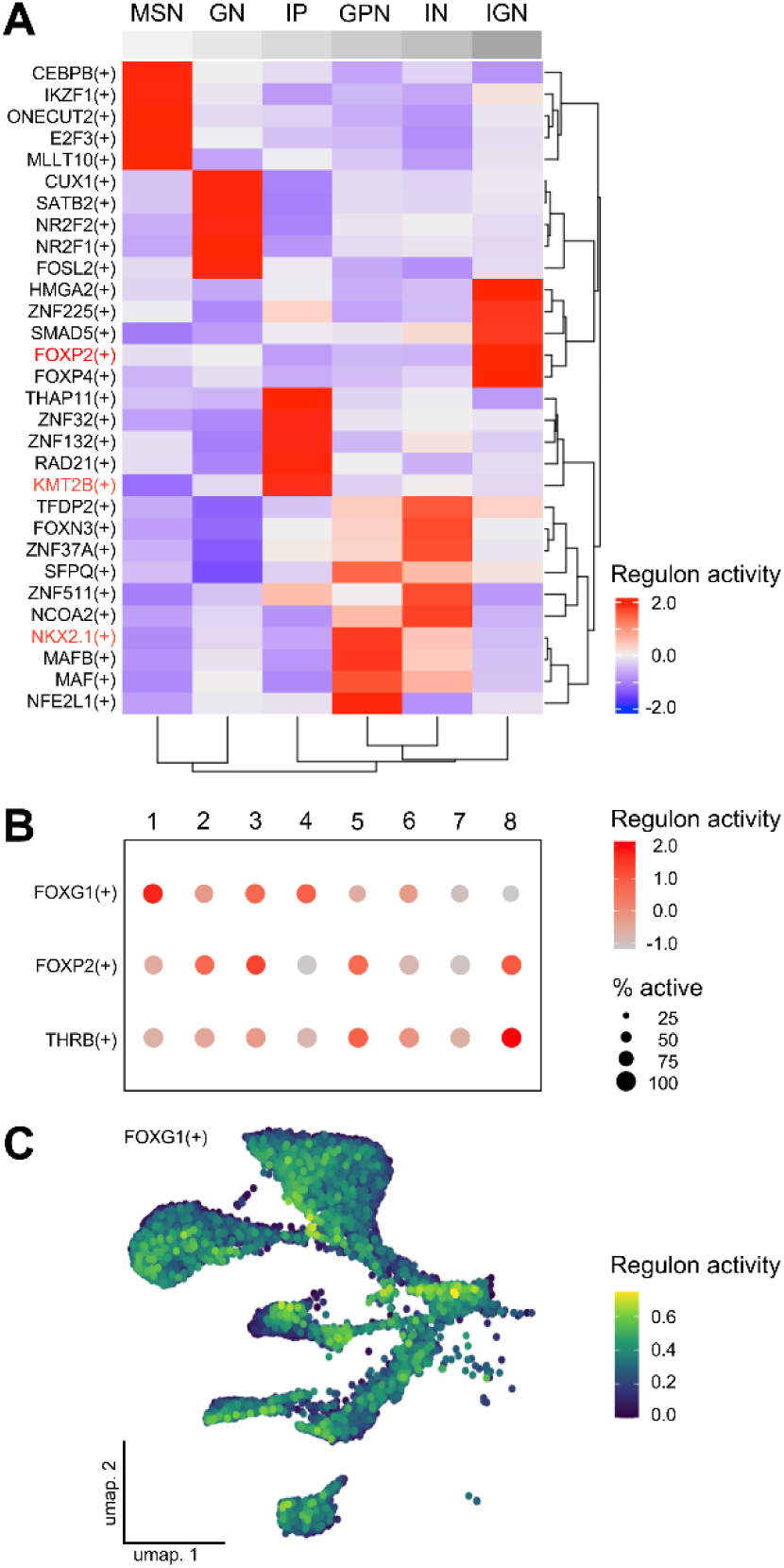
Gene-regulatory networks in basal ganglia development. **(A)** SCENIC regulon activity matrix^39^ showing the top active transcription factors and other proteins that regulate gene expression in each cell type. Proteins discussed in the text are highlighted in red. MSN, GABAergic medium spiny neurons; GN, glutamatergic neurons; IP, intermediate progenitors; GPN, GABAergic projection neurons; IN, interneurons; IGN, immature GABAergic neurons. **(B)** Regulon activity, corresponding to transcription factor activity, of three exemplary genes across donors. 1–8: donor #1–8. **(C)** FOXG1 regulon activity in neuronal cell types.

## Discussion

In this study, we present a transcriptomic atlas of the human basal ganglia spanning their prenatal and early postnatal development to adulthood. We provide the first publicly available single-nucleus transcriptome data of the human perinatal basal ganglia (**Figure 5**) and made it accessible on CELLxGENE (https://cellxgene.cziscience.com/collections/3332ad3e-8599-45af-bef8-17f16cf24245).^41^ Our dataset offers a high-resolution map of gene expression across the major striatal and pallidal cell types. This is particularly relevant for subcortical regions such as the basal ganglia, which have historically been less well characterized than cortical structures, but have very recently gained increasing attention in neuroscience research.^15,19–22^ Importantly, data on spatiotemporal expression is critical for the rational design of gene therapies. This refers in particular to the selection of cell type-specific promoters and AAV capsids, key determinants of transgene specificity, as well as tissue/cell type tropism of vector systems, which require precise knowledge of target cell populations and their time resolved gene expression patterns.^42^ Hence, the integration of cellular identities, developmental trajectories, and gene-regulatory network activities through our dataset provides an open-access resource that supports the definition of therapeutic target cells and critical time windows for intervention, especially for gene-based treatment of pediatric movement disorders.

**Figure 5.**
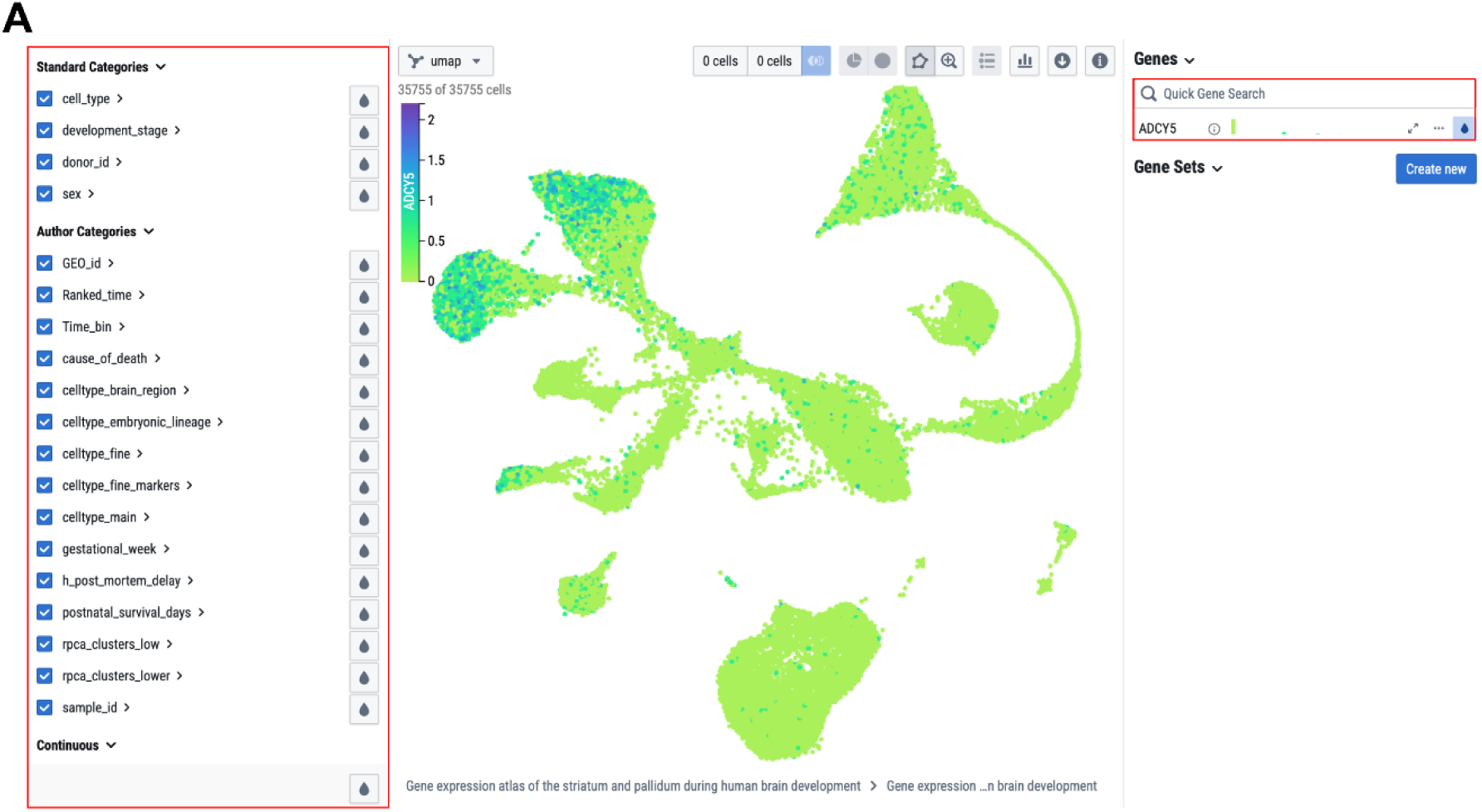
Example application of the publicly accessible CELLxGENE atlas. **(A)** *ADCY5* expression visualized in the UMAP plot of our dataset. Genes of interest can be freely queried using the ‘quick gene search’ function (red box, upper right) and displayed in a color-coded manner across the entire dataset or within selected categories only (red box, left panel).

A key contribution of this study lies in the highlighting of spatiotemporal expression patterns of movement disorder-associated genes from the MDSGene database.^31^ Our findings demonstrate that disease genes differ markedly in both their cellular specificity and the timing of their expression during development, which have important implications for therapeutic design, as illustrated below by four representative use cases:

[1] Patients with heterozygous pathogenic variants in *GNAO1* present with hyperkinetic movement disorders (predominantly dystonia and choreoathetosis, OMIM #617493) and/or epilepsy (OMIM #615473), typically accompanied by developmental delay and intellectual disability.^43^ *GNAO1* encodes the G protein alpha subunit o (Gαo), which is highly expressed in the brain and plays a central role in the regulation of the cAMP signaling cascade, a pathway critical for neuronal processes such as neuromodulation, synaptic plasticity, and excitability.^44,45^ Currently, two gene therapy strategies are being pursued: (**i**) intraparenchymal AAV9-based gene delivery^33^ and (**ii**) a systemic intravenous AAV9-mediated RNA interference (RNAi) approach, aimed at silencing the dominant-negative effect of the hotspot c.607G>A mutation.^30^ Both approaches define the *Striatum* as their primary target region, largely based on expression data derived from rodent models.^33,46^ In contrast, our human dataset demonstrates that *GNAO1* expression is not restricted to the MSN of the *Striatum*, but is also present in projection neurons of the *Globus pallidus* as well as in non-neuronal cell types of the basal ganglia, including oligodendrocytes. These findings suggest that therapeutic targeting strategies may need to be refined to more accurately reflect the broader cellular and regional *GNAO1* expression in the human basal ganglia. As an alternative approach to intraparenchymal delivery, which relies on diffusion and is therefore restricted to single circumscribed brain regions, magnetic resonance-guided focused ultrasound has recently emerged as a minimally invasive strategy to transiently open the blood-brain barrier at predefined locations using low-intensity ultrasound. This approach enables better localized and tissue-specific delivery of systemically administered therapeutics to deep brain structures such as the basal ganglia,^47^ and may therefore represent a potential complementary strategy in this context.

[2] We identified additional species-specific differences in the expression of *ADCY5*. The adenylyl cyclase type 5 (ADCY5), like GNAO1, is a key component of neuronal GPCR-mediated cAMP signaling. ADCY5 catalyzes the conversion of adenosine triphosphate (ATP) to cAMP and represents the predominant adenylyl cyclase isoform in striatal MSN, where it integrates dopaminergic signaling. Patients with dominant pathogenic gain-of-function variants in *ADCY5* present with a broad spectrum of continuous and/or paroxysmal hyperkinetic movement disorders, most commonly chorea, dystonia, and myoclonus (OMIM #606703).^35^ Notably, an unexpected clinical observation in an affected family revealed that caffeine markedly improved motor symptoms.^37^ This effect was subsequently confirmed in a retrospective cohort, in which 87% of participants (26/30) experienced a clinically meaningful improvement, defined as a reduction in overall movement disorder severity of 40% or more.^48^ Mechanistically, caffeine is thought to exert its therapeutic effect *via* antagonism of the adenosine A2A receptor (ADORA2A), which itself stimulates ADCY5.^35,49^ In our dataset, *ADORA2A* expression was restricted to *DRD2*-positive and was not detected in *DRD1*-positive MSN (**Figure S2A**). Combined with rodent studies, which have suggested *ADCY5* expression to be confined to *DRD1*-positive MSN, this would imply that, at least during development, *ADORA2A* and *ADCY5* are not co-expressed in the same cell type. As a consequence, this would not explain the therapeutic effect of caffeine in ADCY5-associated movement disorders.^49^ However, contrary to previous reports, our human data show that *ADCY5* is expressed in both *DRD1*- and *DRD2*-positive MSN. This expanded expression pattern provides a potential molecular explanation for the clinical effects of caffeine, as well as for the partial therapeutic response, because not all relevant cell types may be susceptible to ADORA2A blockade. Taken together, these findings underscore the importance of human-specific reference datasets for understanding disease mechanisms and for guiding the development of targeted therapeutic strategies.

[3] With respect to temporal stratification of gene expression, we identified distinct patterns that delineate therapeutic windows. *FOXG1*, a transcription factor regulating forebrain development, showed predominantly prenatal expression with rapidly declining postnatal expression and regulon activity. These findings suggest that therapeutic interventions targeting FOXG1 might need to be applied very early, potentially even prenatally, in order to be efficient. Patients with dominant pathogenic variants in *FOXG1* develop the FOXG1 syndrome, also called the congenital variant of Rett syndrome (OMIM# 613454). This disease is characterized by moderate-to-profound global developmental delay, hyperkinetic movement disorders, axial hypotonia, psychiatric manifestations, epilepsy, and spasticity.^50^ In a heterozygous *Foxg1* mouse model, early postnatal (postnatal day P1) intracerebroventricular administration of an AAV9-*Foxg1* based gene therapy partially rescued structural brain abnormalities, including *Corpus callosum* dysgenesis, dentate gyrus malformations, and myelination abnormalities by the age of P15 and P22.^29^ Such abnormalities had also been observed in neuroimaging and neuropathological studies of human patients.^51^ Nevertheless, it remains unclear to what extent these findings could be translated to humans. For example, in the mouse, corticogenesis is still ongoing at birth, whereas in humans corticogenesis has been completed for approximately 90 days by the time of birth.^52^ Direct interspecies comparisons for basal ganglia development are lacking, but taking the trajectory of overall brain development as a proxy, P1 in the mouse corresponds to postconceptional day 141 (20 weeks-of-gestation) in humans.^52^ It is therefore important to ponder whether immediate postnatal gene therapy in mice, whose neurodevelopmental state at birth is comparatively immature, would be a good preclinical model for a postnatal intervention in human neonates. Based on our data, it appears possible that the therapeutic window in humans may not extend substantially into postnatal stages. On the other hand, it remains possible that the critical window for intervention may be altered or extended beyond the normal physiological expression under pathophysiological conditions. These questions have to be addressed in appropriate preclinical models prior to clinical testing in children.

[4] We observed that *HTT* is expressed during prenatal stages. Patients with heterozygous CAG trinucleotide repeat expansions in *HTT* develop Huntington’s disease (HD, OMIM# 143100), a progressive neurodegenerative disorder characterized by motor, cognitive, and psychiatric symptoms. HD typically manifests in adults and, less frequently, in adolescents depending on the repeat length, which may increase in successive generations (anticipation).^53^ Emerging evidence suggests that HD may also have a developmental component, supported by recent findings of altered cortical development in human fetal HD samples.^54^ In addition, Bocchi *et al.* identified HTT as a major upstream regulator of neurodevelopment in the GE.^14^ Combined with our data, these observations raise the possibility that the disease process begins long before the manifestation of clinical symptoms. Currently, a phase I/II clinical trial is evaluating an AAV5-based gene therapy employing a microRNA approach to silence the *HTT* mRNA.^55^ In a small cohort of 12 patients, this strategy led to a reduction of HD symptoms by up to 75% over a three-year period as compared to controls.^55^ Notably, inclusion criteria required clinically manifest disease.^55^ However, if HD does indeed involve an irreversible neurodevelopmental component, this raises important questions regarding the optimum timing of diagnostics and therapeutic intervention. Given the near-complete penetrance associated with CAG repeat lengths over 40, one could consider whether predictive genetic testing in asymptomatic at-risk minors might be justified to enable timely treatment still in the presymptomatic phase in the future.

In summary, our study provides a comprehensive single-nucleus transcriptomic atlas of the developing human basal ganglia with direct implications for the understanding of pathophysiology and treatment options for pediatric movement disorders. By linking cell type-specific gene expression and regulatory activity to respective stages of human brain development, we establish a framework for defining target cell populations and therapeutic windows. Our open-access resource is expected to support the rational design of gene-based therapies and highlights the importance of integrating human developmental data into translational neuroscience.

## Materials and methods

### Sample collection

Basal ganglia samples (*Striatum* and, in some cases, *Globus pallidus* and *Nucleus accumbens*) from seven neurotypical donors (second trimester to 4.0 postnatal months) were obtained from the NIH NeuroBioBank (**Table 1**). In addition, striatal tissue from one adult female donor (∼60 years) was obtained from the Department of Neuropathology at Charité – University Medical Center Berlin. Brain regions were anatomically assigned by neuropathologists, freshly frozen in isopentane on dry ice, vacuum-sealed, and stored at −80 °C. To ensure sample integrity, only tissue with a postmortem interval of below 27 hours was included. Preterm-born infants survived for a maximum of 24 hours after birth, allowing us to approximate the developmental stage by the gestational age (calculated from the first day of the last menstrual cycle) without the need for age correction. The sex distribution was balanced across donors.

The NIH had received all necessary positive votes from the competent ethics committees, institutional review board, and properly signed informed consent forms from the donors or their representatives. At the Charité – University Medical Center Berlin, approval by the local IRB (Ethikkommission der Charité Campus Mitte; EA1/76/20) and written informed consent of the subject had been obtained as a precondition to use the material in our investigation.

### Sample preparation

Nuclei were extracted following the 10x Genomics protocol for complex tissue (CG000375, protocol rev. C). Fresh frozen brain tissue samples (**Table 1**) were cut into small pieces and immediately transferred into chilled NP40 lysis buffer (10 mM Tris-HCl pH 7.5, 10 mM NaCl, 3 mM MgCl_2_, 0.1% IGEPAL CA-630, 1 mM DTT, 1 U/µL Protector RNase inhibitor (Roche *via* Merck, 3335402001)) containing detergents to lyse cell membranes while preserving nuclear integrity for 5 minutes. During the incubation, samples were homogenized using a pellet pestle on ice. Subsequently, samples were filtered through a 70 µm cell strainer. Nuclei were then pelleted by centrifugation at 500 x *g* for 5 minutes at 4°C, and resuspended in PBS containing 1% BSA and 1 U/μL Protector RNase inhibitor. The last two steps were repeated. The nuclei were then stained with 7AAD in PBS containing 1% BSA and 1 U/μL Protector RNase inhibitor and sorted according to the 647 nm emission signal with a Cytek Aurora (Cytek Biosciences, RRID:SCR_019826) or FACSAria II (BD Biosciences, RRID:SCR_018934). Sorted nuclei were centrifuged at 500 x *g* for 5 minutes at 4°C. The pellet was resuspended in Lysis Buffer (10 mM Tris-HCl pH 7.5, 10 mM NaCl, 3 mM MgCl_2_, 0.01% Tween-20, 0.01% IGEPAL CA-630, 0.001% Digitonin, 1% BSA, 1 mM DTT, 1 U/µL Protector RNase inhibitor) and incubated on ice for 2 minutes to permeabilize the nuclei. Nuclei were then washed with Wash Buffer (10 mM Tris-HCl pH 7.5, 10 mM NaCl, 3 mM MgCl_2_, 1% BSA, 0.1% Tween-20, 1 mM DTT, 1 U/µL Protector RNase inhibitor), centrifuged again, and resuspended in an appropriate volume of Diluted Nuclei Buffer (1X nuclei buffer (from the Chromium Next GEM Single Cell Multiome ATAC + Gene Expression kit (10x Genomics, PN-1000285)), 1 mM DTT, 1 U/µL Protector RNase inhibitor) based on estimated yield and concentration, assuming a loss of approximately 80% of nuclei during lysis. Trypan Blue staining was used to assess nuclei quality and concentration using a hemocytometer. Final concentrations were adjusted to ∼8,060 nuclei/µL by diluting the nuclei suspension accordingly. Nuclei were kept on ice throughout the procedure to preserve integrity.

### Single-nuclei library generation

Libraries were constructed using the Chromium Next GEM Single Cell Multiome ATAC + Gene Expression (GEX) kit (10x Genomics, PN-1000285, PN-1000230, PN-1000212, PN-1000215) according to the manufacturer’s instructions (10x Genomics, CG000338, protocol rev. F). Briefly, open chromatin was tagmented and gel beads-in-emulsion (GEMs) were generated from nuclei, master mix, and gel beads. Subsequently, mRNAs and ATAC fragments were barcoded and mRNAs reverse transcribed, GEMs were cleaned up, and the products were pre-amplified and cleaned up another time. Library construction was performed separately for both modalities. cDNAs were amplified and cleaned up, followed by fragmentation, end repair, and A-tailing, size selection, adaptor ligation, another cleanup, and a final amplification, followed by a last size selection. For ATAC library preparation, adaptors were ligated during a sample index PCR, followed by size selection. cDNA libraries were pooled and sequenced on a NovaSeq® X Plus instrument using a 10B flow cell in 28+10+10+90 mode to a minimum of 325 million reads per sample. ATAC libraries were pooled and sequenced on a NovaSeq® X Plus instrument using a 25B flow cell in 150+10+24+150 mode to a minimum of 355 million reads per sample. The quality of the ATAC libraries was low, which is why they were excluded from all analyses.

### Bioinformatic analyses

Samples were processed in two batches using similar pipelines. Sequencing quality was assessed using FastQC v0.11.9 and MultiQC v1.6. Cellranger-arc 2.0.2^56^ and Cellranger v7.1.0^57^ were used to map the reads to the human reference genome GRCh38 and to extract count matrices of the first multiome batch and the subsequent RNA-only rerun, respectively. We followed a standard workflow for data preprocessing using the Seurat package v4.3.0 in R v4.0.0.^58^ Quality control preprocessing included filtering out low-quality cells with a threshold of 5*MAD (median absolute deviation) of the following metrics: mitochondrial gene counts percentage, total number of features, total number of GEX counts. Doublets were found and removed with scDblFinder^59^ and ambient RNA was corrected for with SoupX.^60^ **Table S1** shows the quality control metrics and resulting number of total cells for each sample. Data were normalized and scaled (regressing out % mitochondrial genes), and highly variable genes were calculated using the `SCTransform()` function of the Seurat package. Samples corresponding to different time points and replicates were integrated using the Seurat package and the rPCA method. To annotate cell types, we identified clusters using the `FindClusters()` function (resolutions = 1.0, 0.5, and 0.1) and manually annotated cell types based on cell type-specific markers found through multiple rounds of sub-clustering. Monocle3 v1.3.4 was used to perform pseudotime analysis using standard parameters. Cell clusters identified as ‘Radial Glia’ and ‘OPCs’ were set as root cell types.^61,62^ To study the activity of known transcription factors, SCENIC v12.1 was used.^39^ Specifically, the log normalized matrices of all cells and of only neuronal cells were exported from R into the pySCENIC docker v12.1. Results were then visualized in R.

## Supporting information

Supplemental UMAPs

## Data availability statement

The R codes used to analyze the single-cell dataset are available at https://github.com/eugeniograceffo/Lange_et_al_2026_basal_ganglia (last accessed: June 01, 2026). The paired-end FASTQ files of the snRNA-seq dataset are available through the Gene Expression Omnibus (GEO) database under the accession number GSE301953 (https://www.ncbi.nlm.nih.gov/geo/query/acc.cgi?acc=GSE301953). The preprocessed snRNA-seq data can be explored interactively on CELLxGENE (https://cellxgene.czisci-ence.com/collections/3332ad3e-8599-45af-bef8-17f16cf24245).

## Data Availability

All data produced are available online at https://www.ncbi.nlm.nih.gov/geo/query/acc.cgi?acc=GSE301953 and at https://cellxgene.cziscience.com/collections/3332ad3e-8599-45af-bef8-17f16cf24245

https://cellxgene.cziscience.com/collections/3332ad3e-8599-45af-bef8-17f16cf24245

## Acknowledgements

We thank the patients and their families for their trust and for permitting the use of their samples for this research. We are grateful to the NIH NeuroBioBank for providing the tissue samples. We thank Robert Opitz for discussions on thyroid hormone-related gene expression and Adina Graffunder for assistance with sample transport. We acknowledge the BIH Medical Genomics Core Facility and the Flow Cytometry Core Facility for technical support. The graphical abstract and **Figures 1A** and **1B** were created in BioRender: Schuelke, M. (2026). The publication licenses can be found here: https://BioRender.com/x4bhkud (graphical abstract, modified), https://BioRender.com/i9loef7 (**Figure 1A, Figure 1B**, modified).

## Author contributions

**BKAL**: data curation, formal analysis, investigation, methodology, supervision, writing of the original draft, and editing the original draft. **EG**: data curation, formal analysis, investigation, methodology, software, validation, visualization, and editing the original draft. **WS**: resources and editing the original draft. **HB**: funding acquisition, resources, and editing the original draft. **MS**: conceptualization, funding acquisition, investigation, methodology, project administration, resources, supervision, and editing the original draft. **NMW**: conceptualization, data curation, formal analysis, funding acquisition, investigation, methodology, supervision, visualization, writing of the original draft, and editing the original draft.

## Declaration of interest

None.

## Funding

**MS** was funded by the DFG under the German Excellence Strategy (EXC-2049-390688087) through the NeuroCure Consortium at Charité – University Medical Center Berlin. **BKAL**, **NMW**, and **MS** were supported by the DFG Research Unit 2841 “Beyond the Exome”, **NMW** is participant in the BIH Charité Junior Clinician Scientist Program for Rare funded by the Charité – University Medical Center Berlin, the Berlin Institute of Health at the Charité (BIH), the Alliance4Rare, and the Berliner Sparkassenstiftung Medizin. **NMW** received a separate grant from the Berliner Sparkassenstiftung Medizin. **NMW** and **MS** were supported by the Federal Ministry of Education and Research (Bundesministerium für Bildung und Forschung, BMBF) as part of the German Center for Child and Adolescent Health (DZKJ). The funders had no role in the design of the study, in the collection, analysis, or interpretation of the data, in the writing of the manuscript, or in the decision to publish the results.

## Notes

### Competing Interest Statement

The authors have declared no competing interest.

### Author Declarations

The ethics committee of Charite -University Medical Center Berlin gave ethical approval for this work: Ref EA1/76/20

### Summary of Updates

In the original submission, the images were missing, they have been replace now.

