## Supplemental UMAPs for "A single-nucleus transcriptomic atlas of human basal ganglia during development forwarding diagnosis and therapy of pediatric movement disorders"

### Supplemental material

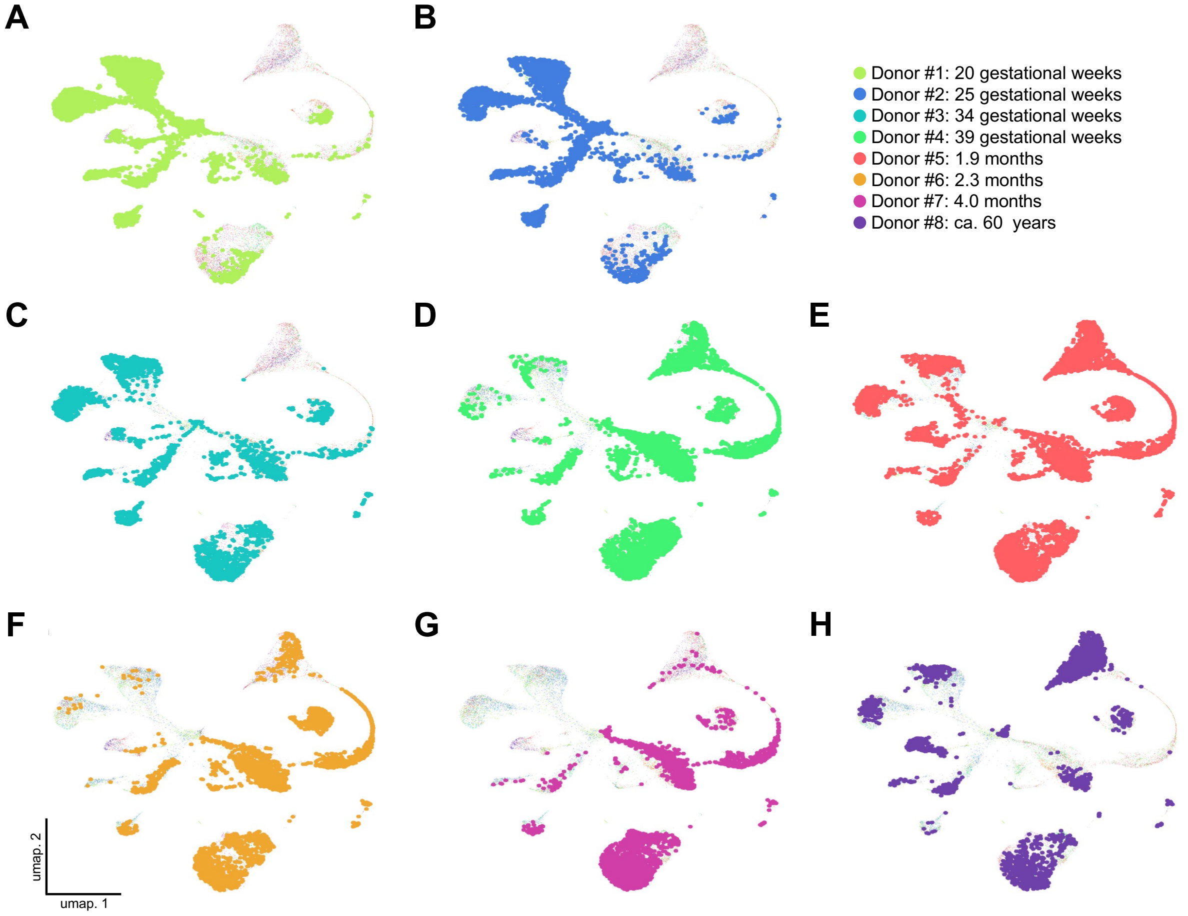


**Figure S1. Cell type taxonomy of brain tissue donors. (A–H)** UMAP plots show the distribution of nuclei from **(A)** donor #1, **(B)** donor #2, **(C)** donor #3, **(D)** donor #4, **(E)** donor #5, **(F)** donor #6, **(G)** donor #7, and **(H)** donor #8 across neuronal and non-neuronal cells.

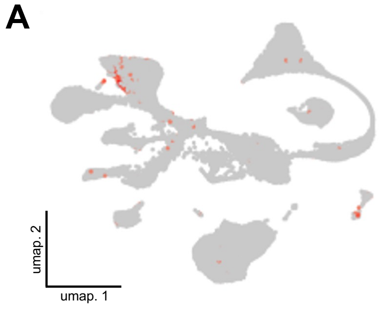


**Figure S2.** ***ADORA2A*** **expression in different cell types. (A)** UMAP plot showing the *ADORA2A* expression in *DRD2*-positive medium spiny neurons.

**Table S1. Quality control metrics and resulting numbers of total cells for each sample.** MADs: median absolute deviations, GEX: gene expression.

| **Sample** | **Number of cells before filtering** | **Cutoff % of mitochondrial genes** | **Cutoff number of features** | **Cutoff number of counts GEX** | **Removed doublet (%)** | **Number of cells after filtering** |
|---|---|---|---|---|---|---|
| Sample_1 | 431 | +- 5 MADs | +- 5 MADs | +- 5 MADs | 16 (3.8%) | 409 |
| Sample_2 | 713 | +- 5 MADs | +- 5 MADs | +- 5 MADs | 56 (8.1%) | 636 |
| Sample_3 | 514 | +- 5 MADs | +- 5 MADs | +- 5 MADs | 33 (6.6%) | 464 |
| Sample_4 | 2814 | +- 5 MADs | +- 5 MADs | +- 5 MADs | 180 (7.1%) | 2364 |
| Sample_5 | 2724 | +- 5 MADs | +- 5 MADs | +- 5 MADs | 148 (5.9%) | 2375 |
| Sample_6 | 1354 | +- 5 MADs | +- 5 MADs | +- 5 MADs | 62 (4.9%) | 1199 |
| Sample_7 | 651 | +- 5 MADs | +- 5 MADs | +- 5 MADs | 26 (4.2%) | 598 |
| Sample_8 | 3626 | +- 5 MADs | +- 5 MADs | +- 5 MADs | 257 (7.4%) | 3197 |
| Sample_1_rep2 | 5793 | +- 5 MADs | +- 5 MADs | +- 5 MADs | 602 (10.7%) | 5023 |
| Sample_2_rep2 | 4195 | +- 5 MADs | +- 5 MADs | +- 5 MADs | 314 (7.7%) | 3777 |
| Sample_3_rep2 | 2964 | +- 5 MADs | +- 5 MADs | +- 5 MADs | 211 (7.3%) | 2693 |
| Sample_4_rep2 | 4692 | +- 5 MADs | +- 5 MADs | +- 5 MADs | 377 (8.8%) | 3910 |
| Sample_5_rep2 | 5406 | +- 5 MADs | +- 5 MADs | +- 5 MADs | 425 (8.4%) | 4627 |
| Sample_6_rep2 | 2622 | +- 5 MADs | +- 5 MADs | +- 5 MADs | 159 (6.5%) | 2300 |
| Sample_7_rep2 | 2708 | +- 5 MADs | +- 5 MADs | +- 5 MADs | 167 (6.5%) | 2403 |
| **Total** | **41207** | | | | | **35975** |
